# Less efficacy of valproic acid monotherapy may be caused by neural excitatory rebound in focal seizures

**DOI:** 10.1101/2021.11.02.21265719

**Authors:** Xiang Zou, Zilu Zhu, Yu Guo, Hongmiao Zhang, Yuchen Liu, Zhengyu Cui, Zunji Ke, Shize Jiang, Yusheng Tong, Zehan Wu, Ying Mao, Liang Chen, Deheng Wang

## Abstract

Valproic acid (VPA) represents one of the most efficient antiepileptic drugs (AEDs) with either general or focal seizures, but a certain percentage of patients are not recovered or even worse, the mechanism under this phenomenon remains unclear. Here, we retrospectively reviewed 16 patients who received awake craniotomy surgery. Intro-operative high density electrocorticogram (ECoG) was used to record the local field potential (LFP) response to VPA treatment. We found the less efficacy of VPA monotherapy was associated with ECoG spectrum power shift from higher to lower frequency after VPA injection, together with increased synchronization of the LFP. Furthermore, we established the computational model to testify the hypothesis that the ineffectivity of VPA may be caused by excitatory dynamic rebound during the inhibitory power increasing. In addition to test the hypothesis, we employed the mice with Kanic Acid (KA)-induced epileptic model to confirm that it would be inhibited by VPA on behavior and neural activity. Also, the neural activity shows significant rebound on spike firing. Then we discovered that the LFP would increase the power spectral density in multiple wave bands after the VPA delivers. These findings suggest that less efficacy of valproic acid monotherapy in focal seizures may be caused by neural excitatory rebound which mediated by elevated inhibitory power.

## Introduction

Epilepsy is one of the most intractable neurological disorders characterized by excessive or hyper-synchronous neurons discharge due to excitatory/inhibitory imbalance ^1, 2^. It is generally assumed that the inhibitory power will reduce the neuron activity. For this reason, the antiepileptic drugs (AEDs) are design as decreasing excitability or increasing inhibitory ability. Valproic acid (VPA) has been introduced in clinical practice over 50 years, and has become the first-line drug for several kinds of epileptic disease nowadays ^3^. Pre- and post-synaptic effects of VPA rely on a broad action spectrum, including the regulation of ionic currents and the facilitation of inhibitory over excitatory neurotransmitter. One of the main mechanisms of VPA is the modulation of GABA-ergic transmission. Specifically, VPA increases the inhibitory activity of GABA by promoting the availability of synaptic GABA ^4^, and enhancing the stimulus-induced responses of both GABA-A and GABA-B receptors ^5^. The selective enhancement of GABA-ergic transmission will increase brain GABA concentration after VPA treatment ^6, 7^. However, VPA monotherapy still has limitations, including inferior efficacy in seizure control in secondary generalized tonic-clonic seizures, focal impaired awareness seizures and some status epilepticus ^8, 9^. In rare conditions, AEDs are reported to aggravate the existing seizures or induce a new seizure type ^10-12^. Besides, VPA-induced seizure aggravation has also been reported in some cases without specific explanations ^13-15^. Despite of the molecular antiepileptic mechanism of VPA, we also need to consider the potential mesoscopic dynamic mechanisms by VPA on neuron population.

The effect of inhibition was long considered to play an important role in EP ^16-18^. Besides, according to our clinic observation, we carefully hypothesized that the inhibitory power from AEDs may have the possibility of inducing membrane potential synchronization, and then cause oscillation of local field potential. In this present study, we found the less efficacy of VPA monotherapy was associated with patients’ ECoG spectrum power shift from higher to lower frequency after VPA injection, together with increased synchronization of the LFP. Then, the computational model which across neuron and population levels on Hindmarsh-Rose neuron model ^19^ was established to testify the hypothesis that the ineffectivity of VPA. Last we performed experiments on mice to verify the hypothesis. On the basis of our results, we propose a relatively universal theory explaining the dichotomous role of inhibitory power.

## Materials and Methods

### Ethics statement

The data collected from human subjects were approved by the Ethics Committee of Huashan Hospital, Fudan University, and was conducted in accordance with the Helsinki Declaration (approval number KY2015-256, KY2019-518). All patients provided written informed consent. All experiments in the study involving living animals were approved by the Institutional Animal Care and Use Committee at Shanghai University of Traditional Chinese Medicine.

### Human subjects’ data collection

We collected data from consecutive 16 patients which 11 of them presented with seizures. As the lesions were related to the eloquent area, all the patients underwent awake craniotomy. In the seizure’s onset patients, 6 of them showed less efficiency in VPA monotherapy pre-operation, and have to receive additional oxcarbazepine regimen post-operation. Four of them developed status epilepticus within 6 hours after surgery and used midazolam maintenance (i.v) for at least 24 hours. The detailed clinical characters were demonstrated in Table 1. During awake craniotomy, high density electrocorticography (ECoG) grid was placed over the lesion and peri-lesion area (8×8 electrode configuration, 5 mm inter-electrode spacing, 1.8-2 mm exposed diameter for each electrode, PMT Corporation, Chanhassen, MN, USA; Beijing Sinovation Medical Technology Corporation, Beijing, China). A 1×6 strip placed distant from the galea served as reference and ground, respectively. We used a Natus Quantum LTM amplifier system (Natus Medical Incorporated, Oakville, ON, Canada) with a sampling rate of 2048 Hz to acquire the ECoG signals. At this sampling rate, we captured signals from 0-878 Hz.

**Table 1.**
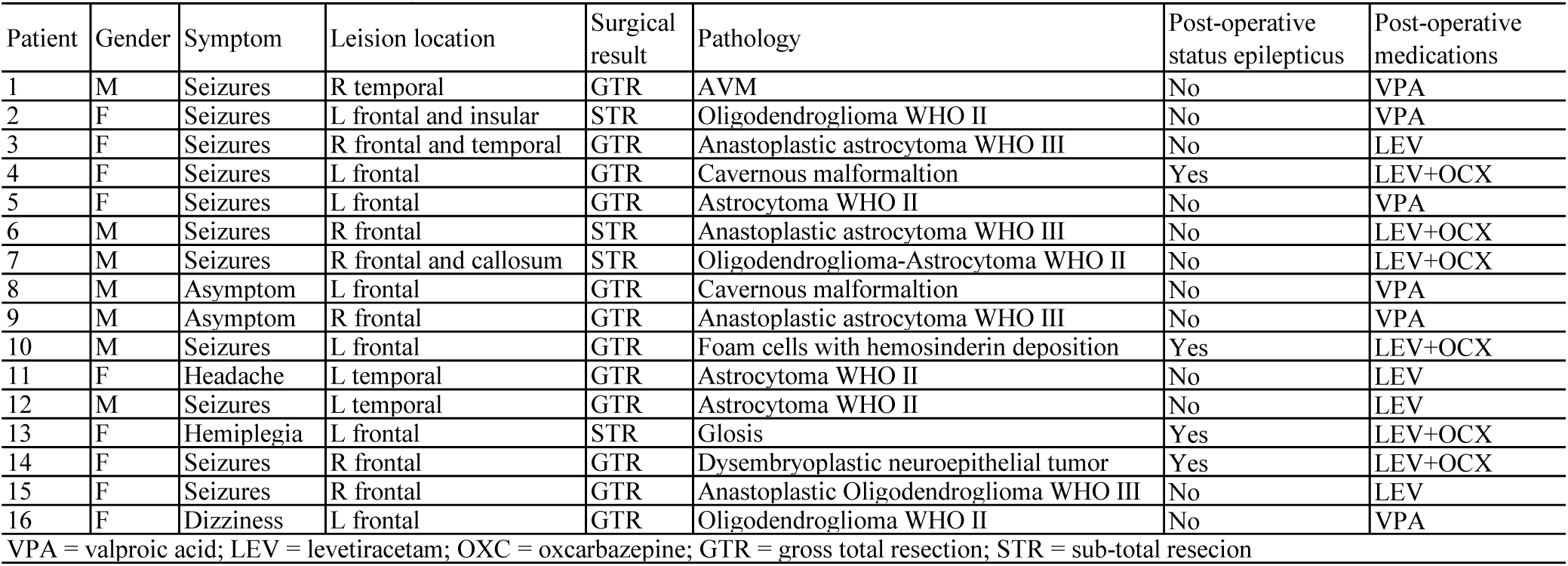
Baseline characteristics of the patients

### Human subject Anesthesia and VPA protocol

We applied an asleep-awake-asleep anesthesia protocol for awake craniotomy. To sedate the patient, we administered the intravenous anesthetics using a dexmedetomidine (DEX) dose of 0.4-0.6 µg/kg of body weight (BW), followed by 0.1-0.2 µg/kg of BW per hour of maintenance infusion. Propofol was infused by target-controlled infusion (TCI) strategy at an effect-site concentration (Ce) of 0.8-1.0 µg/mL along with remifentanil (Ce 0.5 ng/mL). The infusion rate of all three anesthetics was progressively increased until the patient reached a deep sedation stage (Observer’s Assessment of Alertness/Sedation Scale (OAA/S) score of 0-1, which is a measure of consciousness evaluated by an experienced anesthesiologist). Next, we injected a scalp nerve block agent (0.375% ropivacaine + 0.5% lidocain + 1:400000 epinephrine) before securing the patient’s head within a stereotactic Mayfield frame. Then the neurosurgeon would perform the craniotomy when patients were under deep sedation, followed by placing an 8×8 ECoG grid for monitoring. After completing the ECoG placement, we suspended the infusion of propofol and remifentanil. This caused the Ce of propofol to decline and the patients to gradually reverse from their sedation until they reached a moderate sedation stage, i.e., an OAA/S score of 2-3. We continuously evaluated the OAA/S score at intervals of 5 minutes. Once the patient reached an OAA/S score of 5, we considered the patient to be totally awake, and the patients can also perform free talk and limbs movements. When patients were totally awake, the baseline ECoG signal was recorded for 5 minutes before VPA injection. After that, a dose of 0.8g sodium valproate (12-18 mg/kg) was intravenously injected within 10 seconds and ECoG signal was recorded for another 5 minutes.

### Human’s data processing

For the human ECoG signals, time-frequency power spectra analysis was performed as previously described ^20^. Then we calculated the average power during the 5 minutes before and after VPA injection for each electrode. We classified the ECoG frequency spectrum to three frequency bands as slow waves (3-5 Hz), sharp waves (6-12 Hz) and spike waves (13-30 Hz) for each electrode, in accordance with the clinical classification. Power gradient ratio was calculated as difference divided by initial power. Power gradient ratio larger than one is considered to be significant excitatory rebound. Significant excitatory rebound electrodes were summed in each frequency band and each patient for the following analysis.

In addition, we calculated the difference of average power and channels’ synchronization between the two periods. We selected the channels of power shifted as the interesting pairs. And the instantaneous phase of the band signals (filtered) is obtained by Hilbert transform.

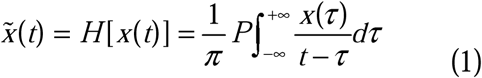

While P is Cauchy value, then the discrete-time analytic signal via Hilbert transform and the instaneous phase value can be computed as below equations

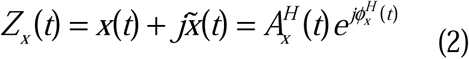

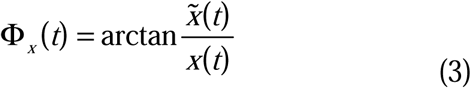

The instaneous phase value for the band signals from another channel can be computed too:

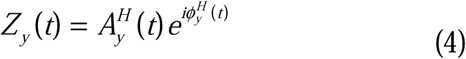

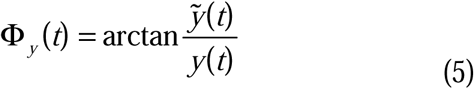

The synchronization between different channels is measured by Phase Synchronization Index (PSI, or phase locking value, plv, ^21^. Then, the phase-locking value, plv) between two signals can be obtained

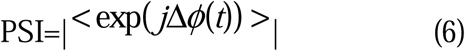

While <.> represent the mean value within certain time windows, Δ*ϕ*(*t*) is the instaneous phase difference between signals x(t) and y(t). The PSI range from 0 to 1, corresponding to synchronization and non- synchronization.

### Statistical Analysis

Except where indicated otherwise, all results are reported as mean standard error of mean. Statistical analysis was performed using the Wilcoxon rank-sum test for nonparametric data, and determined by one-way ANOVA with Bonferroni test for post-hoc analysis. Differences in clinical features between mono/duotherapy as well as no/post-operative status epilepticus groups were assessed using the Fisher exact test. Differences were considered significant for values of p < 0.05. All the data were analyzed using SPSS version 18.0 (IBM Corp., Armonk, New York, USA) and GraphPad Prism 9 software (GraphPad Inc. San Diego, CA, USA).

### KA Induced Epilepsy Animal Models

We use two different doses (the low-dose 10 mg/kg ^22^ and the high-dose of 30 mg/kg ^23^) of KA to induce epilepsy (intraperitoneal injection, i.p.) in separate animal groups. The animal group with low-dose group was chosen due to the low rate of animals’ mortality. An hour behavior monitoring was performed to observe the seizure condition immediately after KA i.p. injection. The epilepsy-like behavior includes wet-dog shaking, forelimb clonus or tonic-clonic convulsions ^24-27^. All behavior data were presented as mean ± s.e.m. Differences were considered significant if *p* value was lower than 0.05.

### VPA control group and antiepileptic animal Models

After 30 minutes free exploring in the home cage, the VPA control group mice were given five times of antiepileptic drug VPA at 70 mg/kg body weight for every two minutes (total in 10 minutes). Then the animals were observed the behavior and the neural activity simultaneously. As the antiepileptic model, the mice were given VPA two minutes after KA injection; the VPA was injected five times at 70 mg/kg body weight for every two minutes and then at 35 mg/kg dose for every 5 minutes until the epilepsy behavior disappear (total in 1 hour). VPA treatment uses continuous i.p. injection of low dose to alleviate the possible side effects of VPA and reduce the animal’s mortality ^28, 29^.

### Animal surgery and *In vivo* recording of neural activity

Male C57BL/6J mice were housed in our University animal facility with a 12 hours light-dark cycle and supplied Food and water ad libitum. All animal procedures were approved by the Shanghai University of Traditional Chinese Medicine Institutional Animal Care and Use Committee. Electrophysiology data were obtained from hippocampal CA1 brain region of freely behaving mice with implantation of 32-channel recording arrays (total 21 mice) ^30, 31^. The recording electrodes consisted of movable bundles of tetrodes, which were constructed by twisting four fine wires (California Fine Wire #100-167). The animals were handled for minimal 3 days prior to electrode implantation surgery. After several days of animal handling, the animal was anesthetized with an i.p. injection of 100 mg/kg pentobarbital sodium. The electrode tips were positioned to the specific brain region above the hippocampal CA1 (AP -2.3 mm, ML 2 mm, DV 1 mm). To eliminate the possible effect of barbiturates on neural activity and allow sufficient recovery time, the mice were placed at homecage for at least 1 week before neural recording.

We recorded the neuronal activity from the hippocampal CA1 region in free behaving mice. The electrode was slowly advanced to the destination region (proximately 35 µm per day). After the *in vivo* recording, the mouse was anesthetized and a certain amount of current was delivered to the electrode tip in order to mark the position of electrode tip. Then the position was confirmed by histological staining (Nissl, 0.1% cresyl violet) of the brain tissue slice.

### Animals’ data collection and processing

The single unit spike and local field potential from mice were simultaneously recorded by an OmniPlex Neural Data Acquisition system (Plexon Inc., Dallas, TX). Spikes which detected at adjustable online thresholds were collected at 40 kHz. Then these spikes were sorted and identified by Offline Sorter 4.0 software and Matlab program (KlustaKwik 1.7 and MClust 3.5: http://redishlab.neuroscience.umn.edu/MClust/MClust.html). Peri-event histograms, correlation with LFP, Coherence of LFP and power spectral density (PSD) were conducted in NeuroExplorer 5 (Nex Technologies). All smooths with histograms were conducted by using a Gaussian filter (filter width = 3 bins).

### Characterization of neuronal responses

To determine whether a recorded unit was responsive to drug injection, we used the i.p. injection time point as time zero to calculate a peri-event histogram using a 30-s bin size. The neuronal activities during 10 minutes before the i.p. injection were used as baseline to determine confidence intervals. To evaluate whether there were significant changes in firing frequency, we first define 80% confidence intervals as a threshold to detect if there was any significant response (firing rate increase or decrease within certain duration after drug injection). Then if the firing rate change reached the 95%, 99.9%, 99.999%, 99.99999% confident intervals, and the duration was longer than five bins, three bins, two bins respectively ^32^. We will consider this unit has a significant neuronal response (increased or decreased).

### Data availability

The data that support the findings of this study are available from Dr.WANG upon reasonable request.

## Results

### Excitatory rebound is a common phenomenon by VPA treatment in human subjects

All the 16 patients completed their procedures successfully with no intro-operative complications. To investigate the ECoG frequency spectrum power shift by VPA treatment, we analyzed ECoG data from all the human subjects. Typical intraoperative procedure was showed in Fig.1A, and ECoG signals were demonstrated in Fig.1B before and after VPA injection. As mentioned above, we classified the ECoG frequency spectrum to three frequency bands as slow waves (3-5 Hz), sharp waves (6-12 Hz) and spike waves (13-30 Hz) for each electrode. The average spectrum power shift of the three bands with VPA injection was demonstrated in Fig.1C in three typical patients as examples. Firstly, we found that all the 16 patients had excitatory rebound by VPA treatment, presented as the increased spectrum power gradient ratio in at least one electrode. Interestingly, in some excitatory rebound electrodes, the increased lower frequency spectrum power seemed to be transferred from higher to lower frequency bands in situ electrodes after VPA injection (Fig.1D, E). To further confirm the synchronization of these waves after VPA treatment, we picked the brown channels in spike wave bands, green channels in sharp wave bands and purple channels in slow wave bands to calculate the phase locking value of these channels. All three wave bands showed significant increase after VPA injection which implied that the VPA could elevate the synchronization of brain waves (Fig.1F-H).

**Fig. 1.**
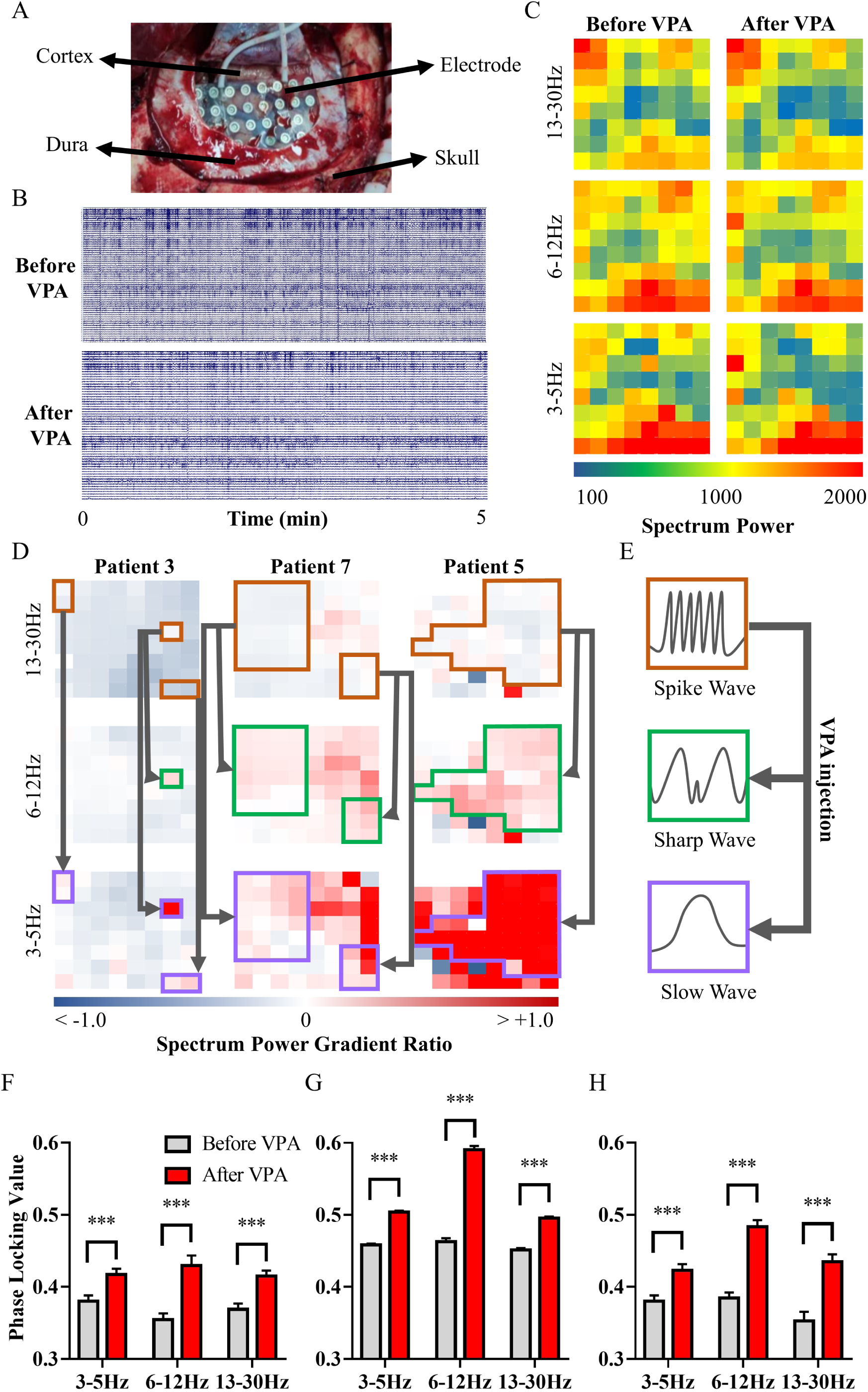
Frequency spectrum power analysis for ECoG signals. **(A)** Intra-operation electrode implantation. **(B)** Typical ECoG signals before and after VPA injection during five minutes. **(C)** Average frequency spectrum power of slow waves (3-5 Hz), sharp waves (6-12 Hz) and spike waves (13-30 Hz) for each electrode, before and after VPA injection during five minutes. **(D)** Spectrum power gradient ratio before and after VPA injection. Brown regions: inhibited spike waves; green regions: increased sharp waves in situ; purple regions: increased slow waves in situ. **(E)** Schematic diagram of spike waves shift to sharp waves and slow waves after VPA injection. **(F-H)** Phase locking value of electrodes of brown, green and purple regions (left: patient 3; middle: patient 7; right: patient 5).

### Significant excitatory rebound induced by VPA is correlated with its ineffectiveness

Clinical characteristics of the 16 patients were showed in Table 1. In 6 of the 16 patients, intraoperative ECOG showed wide significant excitatory rebound (more than 5 electrodes) induced by VPA in any of the three frequency bands. Notably, 5 in those 6 patients had to add oxcarbazepine regimen for seizures control after operation (OR = 45, CI: 2.287-885.601, *p* = 0.008). Moreover, 4 in the 6 patients developed status epilepticus within 6 hours after surgery and had to use midazolam maintenance for seizures control at least 24 hours (*p* = 0.008). For the 10 less rebound patients, 9 of them were seizures free by VPA monotherapy (some changed to LEV for hepatic impairment concern). When considering other possible related clinical characters such as onset symptom, lesion location and extent of resection, etc., none had significant correlations with VPA ineffectiveness. Details were demonstrated in Table 2. Further analysis demonstrated that the amount of significant rebound electrodes in duotherapy group is remarkably larger than in monotherapy group for 3-5Hz band (22.50 ± 10.57 vs. 1.30 ± 0.68, *p* = 0.0128, Fig. 2A). Similarly, the amount of significant rebound electrodes in status epilepticus group is much larger than in no status epilepticus group (32.25 ± 13.52 vs. 1.58 ± 0.70, *p* = 0.003, Fig.2B).

**Fig. 2.**
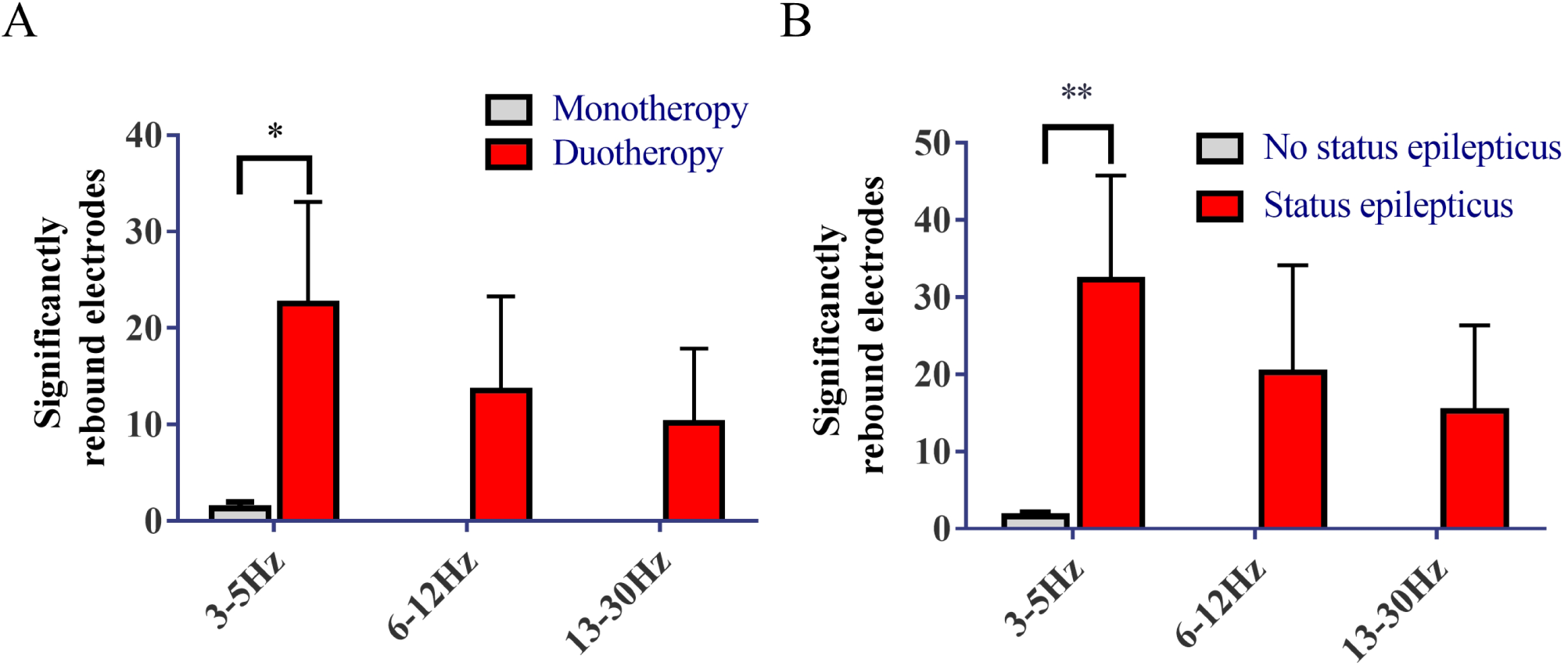
Ineffectiveness of VPA is correlated with significant excitatory rebound. **(A)** Comparison of rebound electrodes amounts between monotherapy and duotherapy group, in three different frequency bands. \**p* < 0.05. **(B)** Comparison of rebound electrodes amounts between status epilepticus and no status epilepticus group, in three different frequency bands. \*\**p* < 0.01.

**Table 2.** Univariate analysis for post-operative antiepileptic regimen and status epilepticus

|  |  | Antiepileptic regimen |  |  | Status epilepticus |  |  |
| --- | --- | --- | --- | --- | --- | --- | --- |
|  |  | Monotherapy<br>(n=10) | Duotherapy (n=6) | *P value* | No<br>(n=12) | Yes<br>(n=4) | *P value* |
| Age |  | 39.2±2.4 | 34.7±4.0 | 0.321 | 39.3±2.0 | 32.3±5.8 | 0.163 |
| Female ratio, % |  | 40% | 50% | 1.000 | 50% | 75% | 0.585 |
| Initial symptom, n (%) |  |  |  | 0.588 |  |  | 1.000 |
|  | Seizures attack | 6 (37.5) | 5 (31.25) |  | 8 (50) | 3 (18.75) |  |
|  | No seizures | 4 (25) | 1 (6.25) |  | 4 (25) | 1 (6.25) |  |
| Hemisphere, n (%) |  |  |  |  |  |  | 0.585 |
|  | Left | 6 (37.5) | 3 (18.75) | 1.000 | 6 (37.5) | 3 (18.75) |  |
|  | Right | 4 (25) | 3 (18.75) |  | 6 (37.5) | 1 (6.25) |  |
| Location, n (%) |  |  |  | 0.234 |  |  | 0.516 |
|  | Frontal | 6 (37.5) | 6 (37.5) |  | 8 (50) | 4 (25) |  |
|  | Temporal | 4 (25) | 0 (0) |  | 4 (25) | 0 (0) |  |
| EOR, n (%) |  |  |  | 0.118 |  |  | 1.000 |
|  | GTR | 9 (56.25) | 3 (18.75) |  | 9 (56.25) | 3 (18.75) |  |
|  | STR | 1 (6.25) | 3 (18.75) |  | 3 (18.75) | 1 (6.25) |  |
| Significant rebound electrodes ≥5, n (%) |  |  |  | 0.008 |  |  | 0.008 |
|  | Yes | 1 (6.25) | 5 (31.25) |  | 2 (12.5) | 4 (25) |  |
|  | No | 9 (56.25) | 1 (6.25) |  | 10 (62.5) | 0 (0) |  |

EOR: extent of resection, GTR: gross total resection, STR: sub-total resection.

### Membrane potential synchronization for excitatory neurons in the neuron cluster with the increasing inhibitory power

The hypothesis that the ineffectivity of VPA may be caused by excitatory dynamic rebound during the inhibitory power increasing was further testified by a computational model we built. In the simulation study, we built a cluster consisted of 800 neurons using the Hindmarsh-Rose model, which was described in Supplementary materials. Generally, this neuron cluster is universal that with variable excitatory/inhibitory neuron proportions, random connections, and variable synapse weights. For each variable situation, we calculated the synchronization factor of membrane potential from the excitatory neurons, in order to represent the entire output projection from the cluster. We adjusted the excitatory/inhibitory neuron proportions from 0.50 to 1.20 (0.02 interval), and normalization inhibitory weight from -1.0 to 0.0 (Gaussian distribution, σ^2^ = 0.25, 0.01 interval). For average synchronization factor on each variable combination, the connections were randomly set a hundred times for calculation. Finally, we got a very detailed 3 dimensional topography of synchronization factor by each variable combination (Fig.3A). Interestingly, the synchronization factor will demonstrate a rebound trend by increasing inhibitory power. In the rebound band, the standard deviation of synchronization factor will also be increased. Fig.3B showed some typical rebound curves in fixed excitatory/inhibitory proportions. Representative LFP and spikes on the synchronization factor curve are showed in Fig.3C, demonstrated the four stages under the increasing inhibitory power. We can see that the spike patterns are almost with high frequency by higher excitatory power in stage 1. When the inhibitory power was increased, some neurons’ spikes are inhibited like in stage 2. In stage 3, the spike frequency became lower but was with more synchronized discharge, resulting in recurrent higher LFP oscillation.

**Fig. 3.**
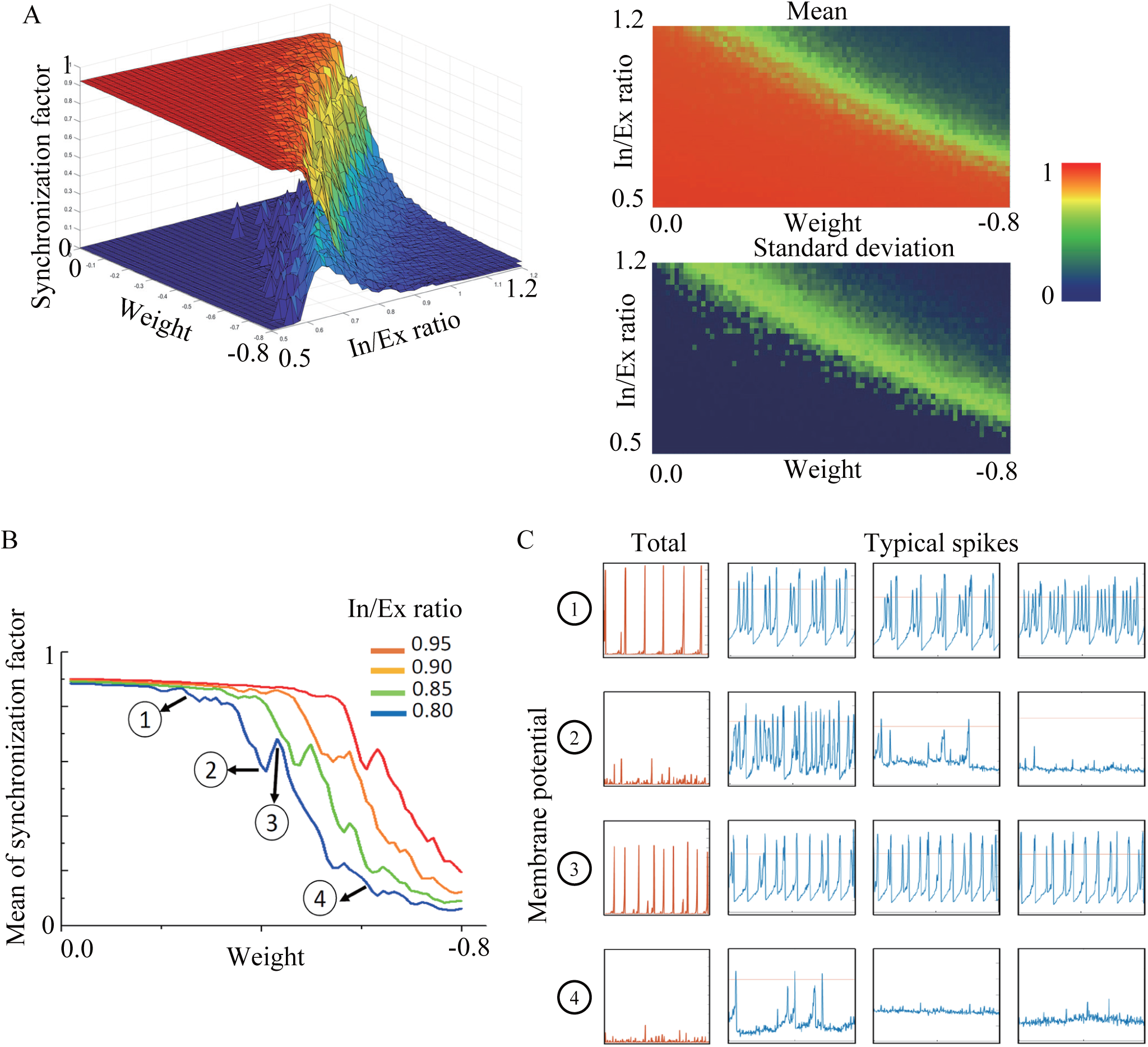
Membrane potential synchronization for excitatory neurons in the neuron cluster with the increasing inhibitory efficiency. **(A)** The value of synchronization factor of membrane potential for excitatory neurons in the neuron cluster, with gradual increasing inhibitory weight and inhibitory neuron distribution. The upper layer in the 3D and the upper panel in the 2D illustration are the mean values, while the lower layer and panel are the standard deviations. The total neuron amount in the cluster is 800, the inhibitory/excitatory ratio varies from 0.5 to 1.2, step 0.02. The mean weight of excitatory synapses is 0.5 (SD = 0.5); while the mean weight of inhibitory synapses varies from -0.8 to 0 (SD = 0.5), step 0.01. Each value is calculated from 50 times independent random connective and assignment patterns, during 10000 sampling units, under each fixed parameters set. **(B)** The value of synchronization factor of membrane potential for excitatory neurons in the neuron cluster, under mean inhibitory weight of -0.5 in different inhibitory neuron distribution. **(C)** Total membrane potential and typical spike patterns of excitatory neurons with the rebounded synchronization factor curve, in four stages ➀-➃. In/Ex: inhibitory/excitatory.

### The seizure induced by KA showed significant suppression by VPA in mice

We employed the mice with Kanic Acid (KA)-induced epileptic model to further test the hypothesis that animal subjects would be inhibited by VPA on behavior and neural activity. A bolus dose of KA with i.p. injection would easily create the models of generalized seizure in freely moving animals. A total of 10 mice were separated into two groups, one was injected KA only and the other was injected KA and VPA (two minutes gap) (Fig.4A). The latency of first epileptic seizure after KA injection was recorded as well as the duration of each epileptic attack. The latency to the first epileptic seizure of KA induced epilepsy modeling mice was 48.8 ± 13.25 s and the KA + VPA treatment group was 69.6 ± 13.34 s. Both groups of animals showed seizure in around 60 s which was ahead of the VPA delivered. No wonder that there was no significant difference in the seizure latency between the KA modeling mice and the KA + VPA treatment group (ns *p* = 0.301) (Fig.4E). The sham group which injected saline instead of KA with VPA did not show any sign of seizure (data did not show). By using Racine Scale, we calculate the seizure stage for every two minutes in the first hour of KA injection. The average stage of VPA treatment was lowered compared to the un-treated group (Fig.4B). Compared to the KA only group, the VPA treatment showed significant suppression during the first 15 minutes (Fig.4C left, \*\*\**p* < 0.001) and 16-45 minutes (Fig.4C middle, \*\*\**p* < 0.001) as well as the 16-45 minutes (Fig.4C right, \*\*\*\**p* < 0.0001). The numbers of seizures in the KA + VPA treatment group within an hour after KA injection was 60.75 ± 18.47 which was significantly lower than KA modeling mice group (167.6 ± 28.47, \**p* < 0.05) (Fig.4D). The duration of each time seizure in the KA modeling mice was 168.8 ± 38.02 s and the KA + VPA treatment group mice was 39.75 ± 6.498 s. The duration of seizure in the KA + VPA treatment group was significantly inhibited (Fig.4F, \**p* < 0.05).

**Fig. 4.**
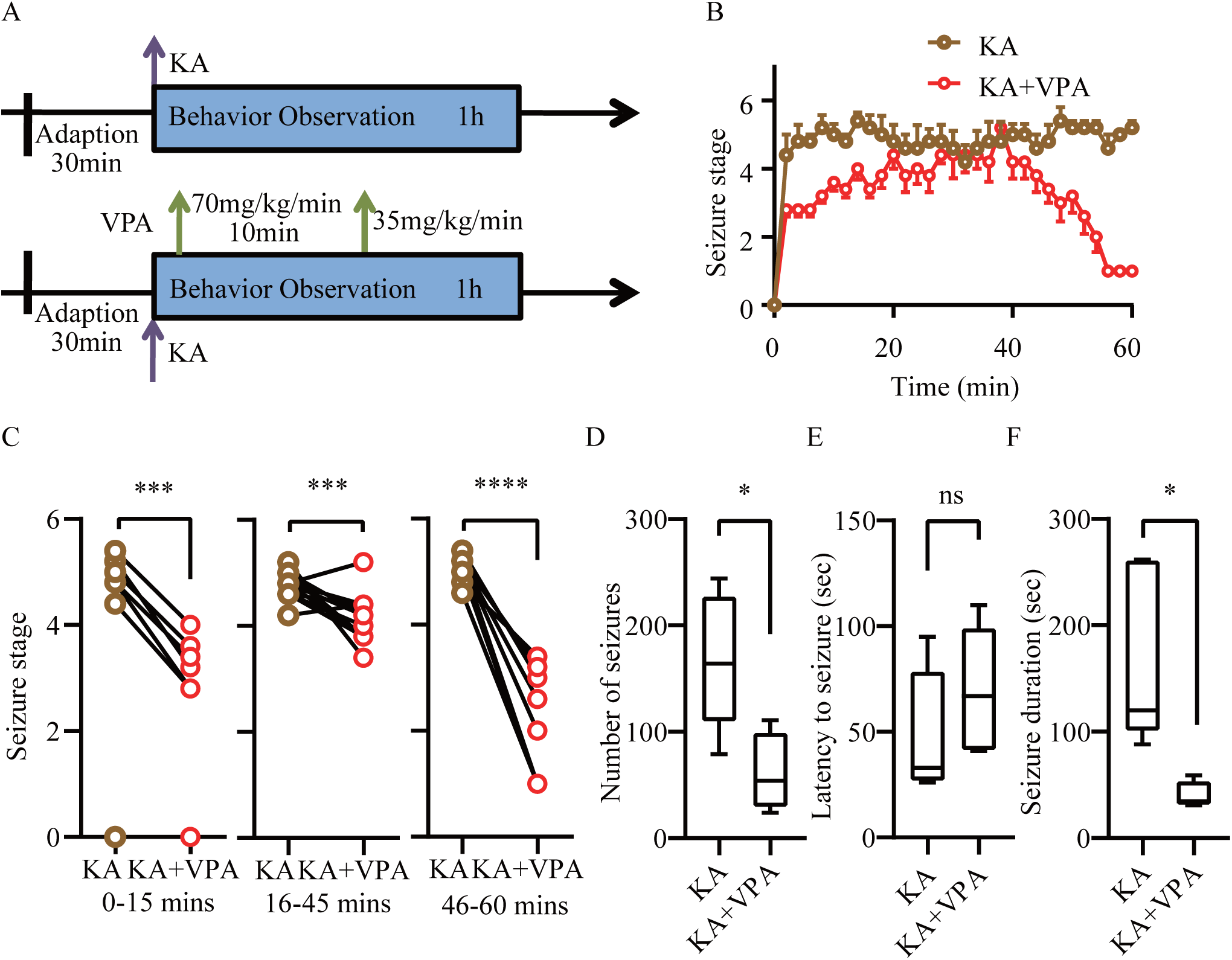
Evaluation of epilepsy behavior in mice. **(A)** Schematic of KA-inducing seizure and KA-inducing with VPA treatment group. **(B)** The seizure stage of KA and KA+VPA groups based on Racine scale. The Racine scale was estimated in every two minutes. **(C)** The seizure stages of KA and KA + VPA group during the first 15 minutes after KA injection (left). The seizure stages of KA and KA + VPA group during the 16-45 minutes after KA injection (middle). The seizure stage of KA and KA + VPA group during the 46-60 minutes after KA injection (right) (Two-tailed paired *t* test \*\*\**p* < 0.001, \*\*\*\**p* < 0.0001). **(D)** The number of seizures during 60 minutes after KA delivered (n = 5 for each group). Data are presented as mean ±SEM (Two-tailed unpaired *t* test \**p* < 0.05,). **(E)** The latency to epileptic seizure. The first seizure discovered after KA injection between 28 to 110 seconds in KA and KA+VPA groups. No seizure was found in VPA group (data did not show) (Two-tailed unpaired *t* test, *p > 0*.*05*). **(F)** The duration of epileptic seizure (Two-tailed unpaired *t* test \**p* < 0.05).

### The neural firing was suppressed with the VPA injection

We employed 64 or 32 channels tetrode micro arrays to record the neuronal spike activity of hippocampal CA1 brain region while the mice freely moving in the home cages (Fig.5A.top). Usually we can monitor several neurons simultaneously from one tetrode such as there were 7 clusters and waveforms recorded from hippocampal CA1 isolated by Offline sorting software (Fig.5A.bottom). We implanted the electrodes in total 7 mice and recorded 139 well isolated neurons in hippocampal CA1 region. The most majority neurons in CA1 brain region which were given VPA injection only showed two different inhibited neuronal spike firing patterns including persistent inhibition until to a plateau (50 units 35.97%, Fig.5B left and Fig.5C) and inhibition with firing rebound (82 units 58.99%, Fig.5B right and Fig.5C). Rest units did not show any firing change to the VPA injection (7 units 5.04%, Fig.5C). Furthermore, we compared the units showing spike rebound with the units of decreased firing or no change in correlation with LFP before or after VPA delivery. No significant difference was found between these three groups of units (Fig.5D left and right). Then we separated the LFP into three different frequency bands (slow wave: 3-5 Hz, sharp wave: 6-12 Hz and spike wave: 13-30 Hz) and calculated the coherence of LFP. All three bands of LFP showed that the coherence was significant lowered after VPA injection (Fig.5E, \*\**p* < 0.01 \*\*\*\**p* < 0.0001).

**Fig. 5.**
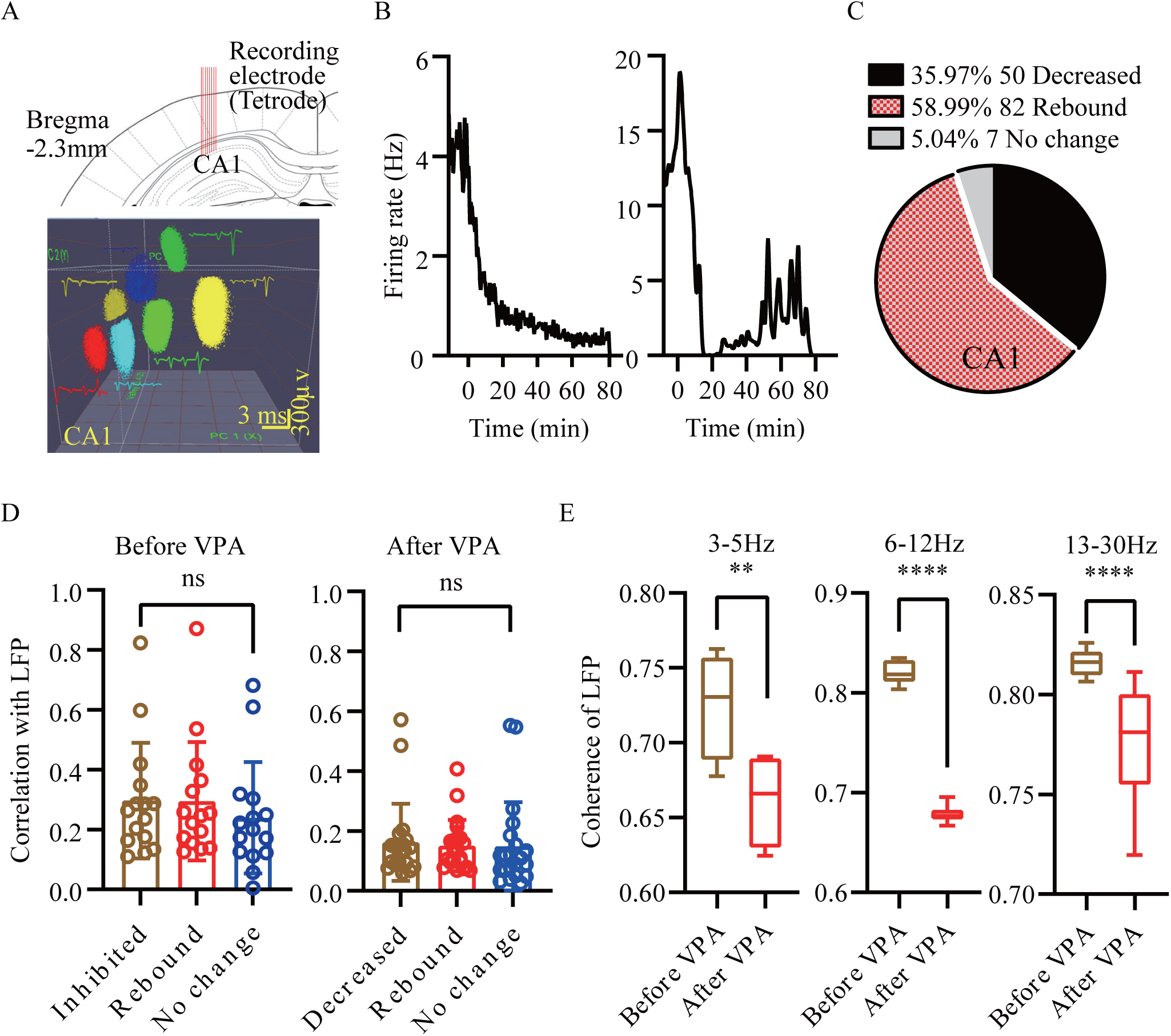
The VPA inhibits the neuron’ firing in hippocampal CA1 area. **(A)** Schematic of tetrode tracks in the mouse hippocampal CA1 brain region (upper), Representative seven well-isolated units (cluster and waveforms) in the PCA plot recorded from the same tetrode in hippocampal CA1 (lower, Scale bar: 3 ms and 300 µv). **(B)** Representative two units from *in-vivo* recording exhibited diversity response to the VPA injection. Peri-event spike histogram show two different responsive units which was an total inhibition (left) and suppressing with an excitation rebound (right). The time zero indicates the time of VPA i.p. injection. **(C)** The ratio of neural activity decreased, rebound and no change units in CA1 brain region. **(D)** The correlation of decreased, rebound and no change units’ spikes with the LFP before and after the VPA injection (Ordinary one-way ANOVA, ns *p >* 0.05). **(E)** The coherence of the LFP before and after the VPA injection displayed in different frequency range (3-5 hz, 6-12 hz and 13-30 hz, Two-tailed paired *t* test, \*\**p* < 0.01, **** *p*< 0.0001).

### The diversity response of neuronal spike activity and LFP in KA modeling mice

We recorded 81 units in total 7 mice in hippocampal CA1 brain region. As we expected, the vast majority units in CA1 regions showed significant response to the KA injection including the inhibition and excitation (Fig.6A and 6C). 77.22% of the responsive units belong to type 2 excitation as well the others were type 1 inhibition (Fig.6C). Also, we could clearly find that the LFP was changed to high amplitude and frequency after KA-inducing (Fig.6B top and bottom). To identify the effect of KA on LFP activity, we calculated the power spectral density and coherence of slow wave, sharp wave and spike wave bands. PSD in the slow wave band (3-5 Hz) was significant increased (\*\*\**p* < 0.001) while the sharp wave (6-12 Hz) shows the same elevation after KA (\**p* < 0.05). Additionally, however, the PSD in spike wave band (13-30 Hz) was reduced significantly (Fig.6D, \*\*\*\**p* < 0.0001). Furthermore, the data of coherence showed that between the slow wave bands, the KA would increase the coherence of LFPs but reduce the coherence between spike wave bands and no statistic significant in sharp wave band (Fig.6E left to right).

**Fig. 6.**
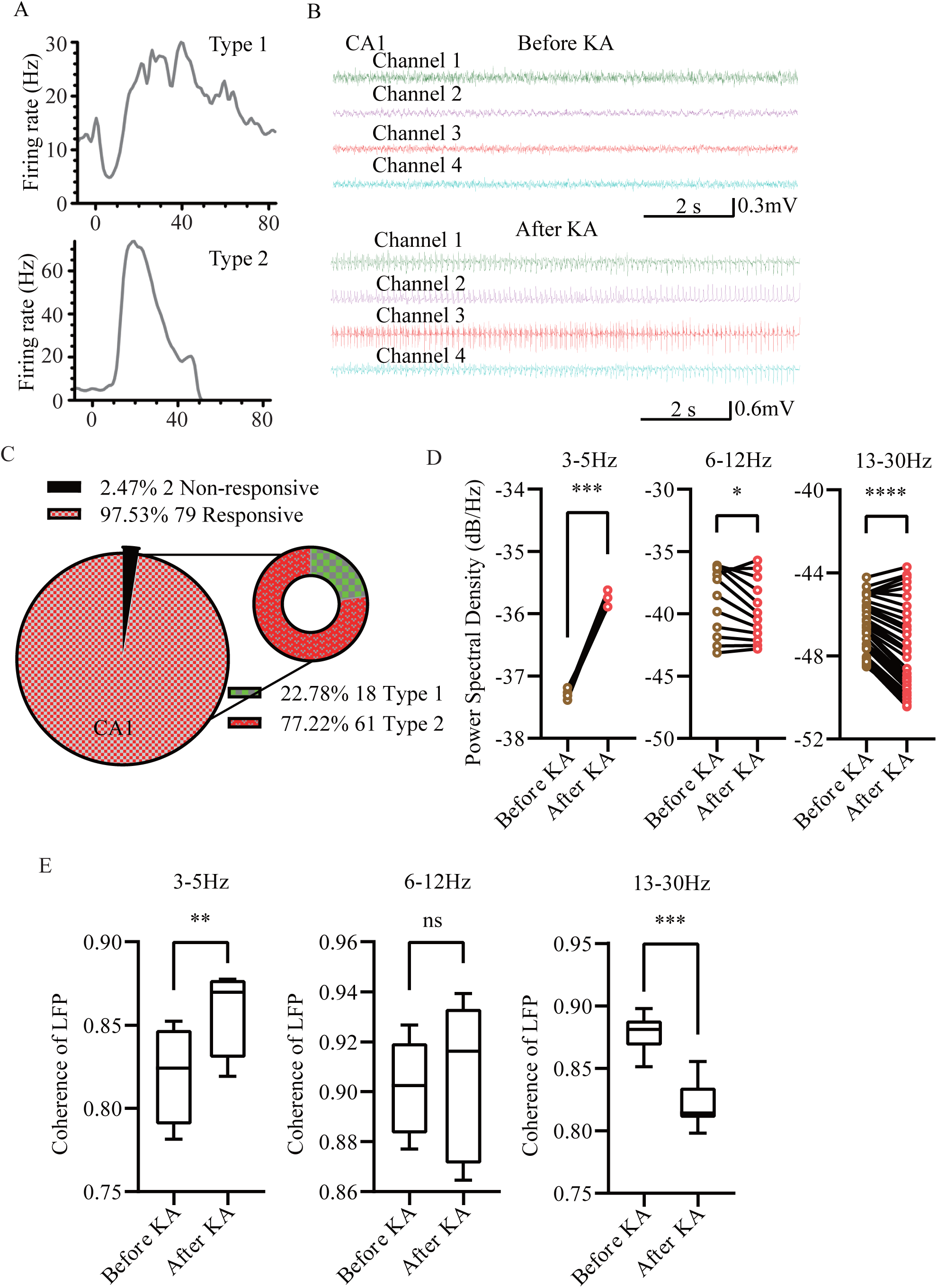
KA-inducing significant increases slow wave band power and reduce spike wave band power in hippocampal CA1. **(A)** Representative units in KA-inducing epileptic animal model. Peri-event spike histogram show two types of responsive units (type 1 and type 2). The time zero indicates the KA i.p. injection. **(B)** Representative LFP traces recorded at different time points before and after KA injection (top: baseline before giving KA, Scale bar: 2 s and 0.3 mV; bottom: after giving KA, Scale bar: 2 s and 0.6 mV). **(C)** The ratio of neural activity show responsive and non-responsive units in CA1 brain region of KA-inducing mice. The percentage of type 1 or 2 of the responsive units at the right panel. **(D)** The power spectral density of different frequency range during baseline recording, after KA injection (left: 3-5 Hz, middle: 6-12 Hz, right: 13-30 Hz, Two-tailed paired *t* test, \**p* < 0.05, \*\*\**p < 0*.*001*, \*\*\*\**p* < 0.0001). **(E)** The coherence of the LFP before and after the KA injection displayed in different frequency range (left: 3-5 Hz, middle: 6-12 Hz, right: 13-30 Hz, Two-tailed paired *t* test, \*\**p* < 0.01, \*\*\**p* < 0.001).

### The neuronal spike activity exhibited rebound excitation after VPA inhibition

After we obtained the neuronal activity results of KA-inducing or VPA-inhibition, we further investigated whether the VPA could effect on the KA-inducing epilepsy. We recorded total 77 units from 7 animals in hippocampal CA1 brain region. There were multiple spike firing patterns we can discover from the different brain regions in KA modeling mice after VPA treatment. We separated the units recorded from CA1 region based on whether the spike neural activity has a rebound phenomenon (rising peak firing after decreasing) (Fig.7A left and middle). There were certain percentages of units exhibited rebound excitation from a few minutes to over 100 minutes after VPA injection. For instance, 53 units 68.83% of CA1 exhibited rebound excitation after VPA injection (Fig.7B). Among these rebound excitation units, we can discover two different firing patterns based on the neural firing properties. The type 1 pattern was increased first after drug delivered and then showed a peak rebound (21 units 39.62% of total rebound units, Fig.7B). The type 2 pattern was decreasing at the beginning and then showed a significant rebound excitation (32 units 60.38% of total rebound units, Fig.7B). Despite these two rebound firing patterns, we discovered other two patterns which showed non-rebound phenomenon. These non-rebound neurons include the units with only one peak of KA-induced firing rate increasing and the non-responsive units (type 3 and type 4). The type 3 showed an increasing firing pattern immediate after the KA injection, which is similar to KA-inducing modeling group (17 units 22.08% of CA1) (Fig.7AB, Fig.6A). Due to the rest units did not have significant response to drug injection, we classified these units as type 4 non-responsive units (7 units 9.09%). For instance, we picked a representative unit of rebound excitation and marked this firing change into 4 stages based on the computational model Fig.3C. The correlation with LFP of four stages showed statistic significant difference between S1 and S2 or S4; S3 and S2 or S4, no significant difference between S1 and S3 (Fig.7C bottom). The results implied that the rebound excitation also correlates to the correlation of spike and LFPs. To identify the effect of VPA on LFP of KA-induced epilepsy model, the PSD analysis was performed for different periods including baseline (no drug injection), after KA-inducing and after VPA. The PSD value after VPA in three wave bands all increased significantly (Fig.7E, *** *p*<0.001). Also, we employed the coherence analysis to calculate these three wavebands coherence of LFP, the results showed that between the slow wave bands, the coherence of LFPs would increase but reduce for the spike wave bands and no statistic significant was found in sharp wave bands (Fig.7F). These findings suggest the power and coherence change caused by VPA injection may explain the phenomenon of rebound excitation.

**Fig. 7.**
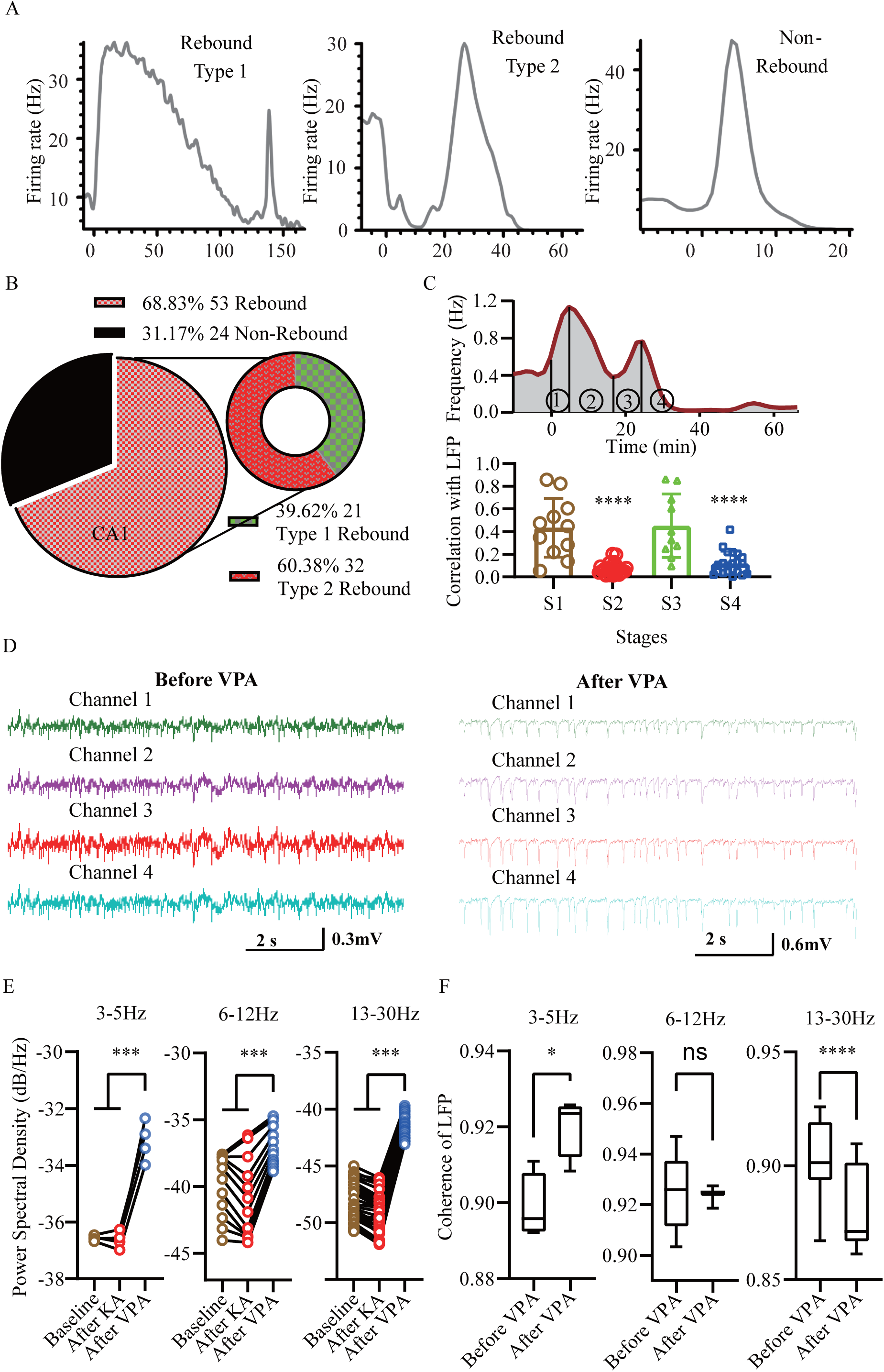
The neuronal activity exhibited rebound excitation after VPA inhibition. **(A)** Representative units from in-vivo recording exhibited diversity response to the KA injection and VPA treatment. The blue dash line indicates the KA i.p. injection. Peri-event spike histogram show that a responsive unit in the VPA suppressed its firing and an excitation rebound later (Rebound type 1 and type 2). The unit without rebound was plotted which exhibited increased firing changes (Non-rebound). The non-responsive unit did not show here. **(B)** The ratio of rebound excitation neural activity by VPA at the left panel and the percentage of type 1 or 2 of the rebound units at the right panel. **(C)** A representative unit with rebound excitation was separated to 4 stages (separated to s1, s2, s3 and s4 by line) based on Fig1.C. The correlation of rebound units’ spikes with the LFP after the VPA injection (Two-tailed unpaired *t* test, \*\*\*\**p* < 0.0001). **(D)** Representative LFP traces recorded at different time points after VPA injection in KA-induced seizure mice (left: before giving VPA; right: after giving VPA). **(E)** The power spectral density of different frequency range during baseline recording, after KA injection and after VPA injection (left: 3-5 Hz, middle: 6-12 Hz, right: 13-30 Hz, Repeated measures One-way ANOVA, \*\*\**p < 0*.*001*). **(F)** The coherence of the LFP before and after the VPA injection displayed in different frequency range (left: 3-5 Hz, middle: 6-12 Hz, right: 13-30 Hz, Two-tailed paired *t* test, \**p* < 0.05, \*\*\*\**p* < 0.0001).

## Discussion

It has long been controversial about the role of IEDs in the transition to seizure, and mutually exclusive studies have demonstrated that IEDs can be either seizure preventing seizure facilitating ^33-35^. In epilepsy patients, although some antiepileptic drug can strengthen GABAergic synapses and inhibit neuronal over-excitation, drug resistance is common in those patients ^36^. Since the mechanism from IEDs to seizures is still unclear, we are also not sure if AEDs will contribute to the transition. According to our findings, the drug resistance may be caused by the generally rebound effect by increasing inhibitory power, even may contribute to the seizure transition. To identify this mechanism from various kinds of patients, a feasible approach is to recognize the LFP spectrum shift from higher to lower frequency high amplitude oscillation. In that case, clinicians should try to increase AEDs amount to cross over the rebound stage before two/three drug regimen or surgical treatment.

Computational models are particularly valuable in the study of epilepsy, which will provide a framework suitable for experimental findings, develop new hypotheses and guide future experiments. To describe the complicated dynamics in epilepsy, various theoretical models have been created to study different spatial scales of a system, from single channels or receptors to neuron population and brain regions. Based on experimental data, very detailed models like multiple neurons with various ion conductance, synapses and connections, models assume neuron population behaviors have been raised for decades ^18, 37-40^. The link between the phenomenological and a network model allows us to gain a deeper understanding of the robustness of the clinical features among patients. Hodgkin-Huxley equations have many parameters and degrees of freedom ^41^, so much more concerns were regard to complicated neuron parameter choices. Here we chose the intermediate Hindmarsh-Rose neuron models, which allowed us to balance neuron parameters and coupling dynamics. In a recent simulation study using the similar model, inhibitory neurons were demonstrated to contribute to spike waves, including interictal spikes in epileptiform activity, which met the *in vivo* recordings ^42^. These simplified neuron models can extract the properties of excitation and inhibition from the electrochemical properties ^19^. Since the excitatory/inhibitory neurons proportion was adjusted by a very wide range of simulation, the rebound phenomenon seems to be universal by the increasing inhibitory power. In addition, the standard deviation of synchronization factor was also increased in the rebound band, indicating the probabilistic nature from IEDs to seizures during this period.

In central nervous system, the unbalanced between excitatory and inhibitory neuronal activity would result in neurological and neuropsychiatric disorders including epilepsy, schizophrenia, autism, and Alzheimer’s disease ^43-46^. The epilepsy is one of the most common neurological disorders of the nervous system and affects 1-2% of the general population, no matter of age or races. Seizures in around 30% patients fail to response to AED due to the complex causes ^47, 48^. To better understand the mechanism underlying the refractory epilepsy, we used the KA-induced epilepsy model of mice to investigate the neurophysiological properties of neuronal activities. The KA is an agonist to activate the glutamate receptor ^49^ while the VPA has been reported to exert anti-epileptogenic effects ^50-52^. In our study, the data of behavior and *in vivo* recording showed the animals have significant epileptic seizure after KA injection. Then the seizure was inhibited by VPA injection. Interestingly, there was significant rebound excitation phenomenon dozens of minutes after drug delivery. Accumulating evidence indicates that bunch of receptors, neurotransmitters increased and other various involved in hyperexcitability in KA-inducing epilepsy ^53, 54^. Since the GABA-ergic interneurons play an important role in the generation of seizures by regulating excitatory-inhibitory balance in the hippocampus. Degeneration of GABA-ergic interneurons would be prevented by the VPA in dentate gyrus during drug-induced status epilepticus ^55^. By elevating the ratio of inhibitory power in excitatory-inhibitory balance, it could be the reason for the rebound excitation after VPA.

There are still some limitations in this study. In the context of epilepsy, the functional properties of the neuron cluster model mainly focus on the dynamic of membrane potential and neuron interactions, which are disrupted by a possibly large set of factors at the cellular and molecular levels. It is therefore needed to sacrifice some biological accuracy to model seizure dynamics in favor of the inhibitory power we concerned about. In human studies, although the result can support our hypothesis of LFP frequency spectrum shift by VPA, the sample size is not that enough. Actually, indications of the awake surgery for epilepsy are very strict, which result in the rarity of patient.

In summary, our findings demonstrate that the increasing inhibitory power may generally contribute to neuron synchronization and LFP frequency spectrum shift, which will induce the epileptic recurrence during AEDs treatment. This study can provide a better understanding of the pathological changes in epilepsy and the potential medication regimen for epilepsy control.

## Data Availability

All data produced in the present study are available upon reasonable request to the authors.

## Abbreviations

AEDs: Antiepileptic Drugs
KA: Kanic Acid
VPA: sodium Valproate
LFP: Local Field Potential
i.p.: intraperitoneal
ECoG: electrocorticography
PSD: Power Spectral Density
DEX: dexmedetomidine
TCI: Target-Controlled Infusion
OAA/S: Observer’s Assessment of Alertness/Sedation Scale
IEDs: Interictal Epileptiform Discharges
PSI: Phase Synchronization Index
PLV: Phase Locking Value

## Acknowledgements

We are indebted to the epilepsy patients and their relatives for their generous assistance with our research.

