## Supplemental file for "Less efficacy of valproic acid monotherapy may be caused by neural excitatory rebound in focal seizures"

**Supplementary files**

### 1. Materials and Methods

In order to reveal the relationship between the excitatory/inhibitory power in a neural cluster, and its membrane potential synchronization, we conduct the simulation experiment based on the Hindmarsh-Rose model (Supplemental Fig.1.A). Specifically, given the total number of neurons and the distribution parameters of excitatory neurons fixed, we study the effect brought by inhibitory neuron parameters, on the synchronization factor $R$ of excitatory neurons. Our experiment is conducted on MATLAB R2019, on a PC with an Intel i7-7700K CPU and 16 GB RAM.

#### 1.1. Theoretical formulation

In this experiment, we adopt the single neuron Hindmarsh-Rose model (Hindmarsh and Rose, 1984) as the formularized spiking-bursting behavior pattern for the neurons' membrane potential. In order to facilitate the MATLAB realization, the model expression is simplified as follows:

$$\begin{cases} \dot{x}(t) = y(t) + 3x(t)^2 - x(t)^3 - z(t) + I(t) \\ \dot{y}(t) = 1 - 5x(t)^2 - y(t) \\ \dot{z}(t) = \mu(4(x(t) - x_0) - z(t)) \end{cases} \quad (1)$$

where $t$ indicates time stamp, $x$ stands for neurons' membrane potential, $y$ and $z$ are the variables account for the status of ion transport across the membrane through the ion channels. $u$ and $x_0$ are non-related constant. $I$ represents the external electrical stimulation of the neurons. Supposed that the status of $x(0)$, $y(0)$ and $z(0)$ are initialized, then the $x(t)$, $y(t)$ and $z(t)$ can be solved via iteration of Equation 1, 2:

$$\begin{cases} x(t+1) = x(t) + \dot{x}(t) \times 1 \\ y(t+1) = y(t) + \dot{y}(t) \times 1 \\ z(t+1) = z(t) + \dot{z}(t) \times 1 \end{cases} \quad (2)$$

We name an execution to be a "simulation round", where the Equation 1 and 2 are calculated by iteration with $t$ going from 0 to $T$. In one simulation round, we can get the membrane potential of all the neurons at each time stamp from 0 to $T$. Then we can derive statistics describing the developing progress of the membrane potential over a time interval shorter than or equal to $T$. The statistics can be such as the mean, standard deviation or the synchronization factor of the neurons' membrane potential.

As for the the synchronization factor *R*, the calculation of the synchronization factor $R_{(m,T)}$ within a time stamp interval *[m,T]* during one simulation round, is defined as the following:

$$\overline{V_t} = \frac{1}{N}\sum_{i=1}^{N} V_{i,t} \qquad (3)$$

$$R_{(m,T)} = \frac{\frac{\sum_{t=m}^{T}\overline{V}_t^2}{T-m+1} - \left(\frac{\sum_{t=m}^{T}\overline{V}_t}{T-m+1}\right)^2}{\frac{1}{N}\sum_{i=1}^{N}\left(\frac{\sum_{t=m}^{T}V_{i,t}^2}{T-m+1} - \left(\frac{\sum_{t=m}^{T}V_{i,t}}{T-m+1}\right)^2\right)} \qquad (4)$$

where t indicate the time stamp ranging from m to *T*, *N* is the number of the excitatory neurons, $V_{i,t}$ is the membrane potential of the $i^{th}$ neuron at the time stamp of t, i.e. $x_i(t)$.

### 1.2. Experiment design

To reveal the developing progress of cranial membrane potential, we study the correlation between inhibitory neuron parameters and synchronization factor *R* of excitatory neurons. There are mainly two independent variables to control and observe in this study, i.e. *D* representing the ratio of the inhibitory and the excitatory neurons, and *u* representing the mean value of inhibitory neurons' membrane potential. Therefore our program simulation is to grid search these two independent variables, and to analyze the mean and standard deviation of the dependent variable, i.e. the synchronization factor *R*.

Specifically, we traverse *D* from 0.6 to 3, with the step of 0.02, and *u* from 0 to 1, with the step of 0.02. The total grid search number is 120 × 50 = 6,000. Within each particular grid of *D* and *u*, 50 round of simulations are conducted, to obtain the mean and standard deviation of $\{R^i_{(5,000,10,000)} | 1 \leq i \leq 50\}$, i.e. $U_{R_{(5,000,10,000)}}$ and $\delta_{R_{(5,000,10,000)}}$. During one stimulation round *i*, as is shown in Figure 2, Equation 1, 2 are computed iteratively, while the temporal variant parameter (instead of the constant) in Equation 1 are saved separately in the relative cell arrays with the index of time stamp *t*. Then, the Equation 3 and 4 are computed to obtain $R^i_{(5,000,10,000)}$ with the cell array of the *X* over time *t*. The time stamp interval is chosen to be [5,000, 10,000] in pursuit of distribution stability. Therefore, the total simulations rounds are 120 × 50 × 50 = 300,000.

The detailed execution and workflow of our experiment is illustrated in the flowchart in Figure Supplemental Fig.1.B, and the important stimulation round is depicted in Supplemental Fig.1.C.

### 1.3. Data structure and parameter initialization

In terms of the data structure of our simulation algorithm, we simplify the mutual neural connection pattern, so that each neuron has time-invariant (constant) mutual connections towards all the other neurons. Therefore the data structure of the neuron connections is designed to be a directed complete graph, which is represented as a constant square matrix $W_{N\times N}$ with a diagonal of zero. Element $w_{ij}$ in $W$ indicates the weight from the $i^{th}$ neuron towards the $j^{th}$ neuron. The membrane potential, $x$ in Equation 1, is assigned as a vector $X_{1\times N}$, with the same design of $Y_{1\times N}$, $Z_{1\times N}$, constant $\mu_{1\times N}$, constant $X_{01\times N}$ for $y$, $z$, $u$, $x_0$. The external electrical stimulation I in Equation 1 is also a vector which is defined as the following:

$$I_{1\times N} = X \times W \quad (5)$$

In our experiment, we assume that the neuron connection weights and membrane potential are all initialized obeying the Gaussian distribution. The total number of both the excitatory and inhibitory neurons is 400. With $D$ representing the inhibitory and excitatory ratio, there are $\frac{400\times D}{1+D}$ inhibitory neurons and $\frac{400}{1+D}$ excitatory neurons. The membrane potential of the excitatory neurons is randomly initialized by the Gaussian distribution with a mean of -1.6, a standard deviation of 0.2.

The membrane potential of the inhibitory neurons is also randomly initialized by the Gaussian distribution with a mean of -1.6, a standard deviation of 0.2. The weight from the excitatory is initialized by the Gaussian distribution with mean 0.001, standard deviation 3. The weight from the inhibitory is initialized by the Gaussian distribution with mean $u$, standard deviation 3. The initial values of the $y(0)$ and $z(0)$ are 0, and the initial value for $\mu$ is 0. The membrane potential thresh for the definition of a spike is 2.

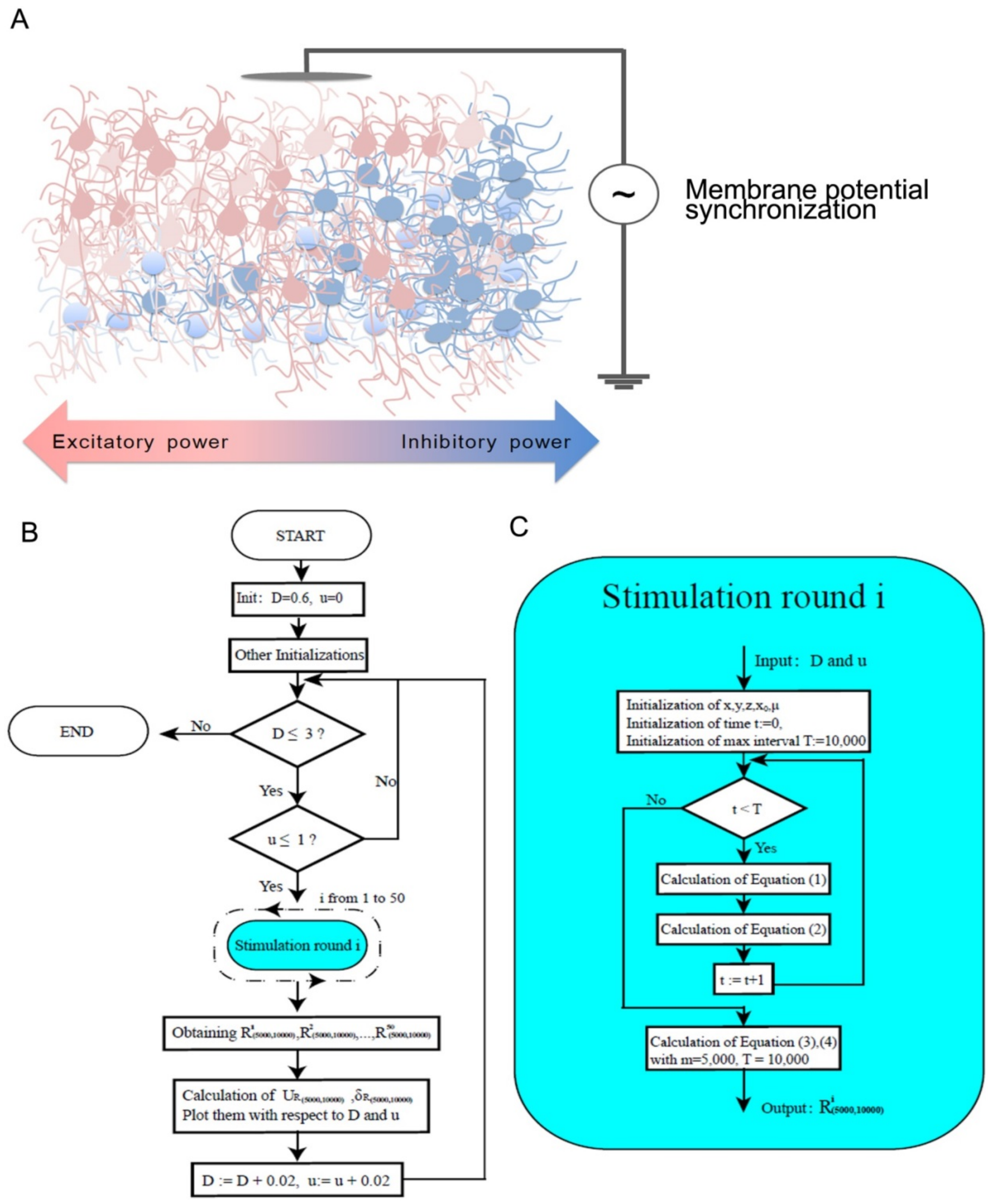


### Figure legends

Supplemental Figure 1. The illustration of the simulation experiment on membrane potential synchronization in a neural cluster.

(A) the main variables in the neural cluster was studied both in the distribution of excitatory/inhibitory neurons, and the average inhibitory weight;

(B) the framework of simulation experiment;

(C) detailed steps in the stimulation round.
